# Exercise-induced symptoms in young childhood cancer survivors

**DOI:** 10.1101/2025.06.16.25329717

**Authors:** Maša Žarković, Carina Nigg, Christina Schindera, Myrofora Goutaki, Sonja Lüer, Marc Ansari, Claudia E Kuehni

**Affiliations:** Institute of Social and Preventive Medicine, University of Bern, Switzerland; Graduate School for Health Sciences, University of Bern, Switzerland; Division of Paediatric Oncology/Haematology, University Children’s Hospital Basel, University of Basel, Switzerland; Division of Paediatric Respiratory Medicine and Allergology, Department of Paediatrics, Inselspital, University Hospital, University of Bern, Switzerland; Division of Pediatric Hematology and Oncology, Department of Pediatrics, Inselspital, Bern University Hospital, University of Bern, Switzerland; Division of Pediatric Oncology and Hematology, Department of Women, Child and Adolescent, University Hospital of Geneva, Switzerland; CANSEARCH Research Platform for Pediatric Oncology and Hematology, Department of Pediatrics, Gynecology and Obstetrics, Faculty of Medicine, University of Geneva, Switzerland

**Author notes:** Corresponding author Claudia Kuehni, Childhood Cancer Research Group, Institute of Social and Preventive Medicine, University of Bern, Mittelstrasse 43, 3012 Bern, Switzerland.

## Abstract

Purpose

Exercise-induced symptoms (EIS) are common in children and understudied in childhood cancer survivors (CCS). We assessed the prevalence of EIS in CCS and identified associated risk factors.

**Methods:** We included children aged 6-20 years who had been diagnosed with cancer ≥ 1 year(s) before study entry, had completed cancer treatment, received pediatric oncology follow-up care, and were treated with systemic anticancer treatment, chest surgery, radiotherapy, or hematopoietic stem cell transplantation. Participants completed a questionnaire on respiratory symptoms and lifestyle. To explore risk factors of EIS, we used multivariable logistic regression and calculated population attributable fractions (PAFs).

**Results:** Of 196 participants (median age 14 years [IQR 10-17]), 46 (24%) reported EIS, including dyspnea (14%), cough (12%), and wheeze (7%). EIS were more common among females (OR 2.5, 95%CI: 1.1-5.9), older survivors (OR 1.2 per year, 95%CI: 1.1-1.3,), and those with obesity (OR 4.7, 95%CI: 1.1-19.6), asthma (OR 10.1, 95%CI: 3.3-31.2), and physical inactivity (OR 2.9, 95%CI: 1.3-6.6). Chest-directed radiotherapy tended also to increase the risk (OR 2.4, 95%CI: 0.6-9.2). Twenty percent of EIS were attributable to asthma, 18% to physical inactivity, and 7% to obesity, with a combined PAF of 44%.

**Conclusions:** EIS affect one in four CCS and are primarily associated with risk factors common in the general population rather than cancer treatments.

Implications for Cancer Survivors

Clinical investigation and management of common causes of EIS—particularly asthma, physical inactivity, and obesity—could reduce symptom burden and support long-term health in CCS.

## Introduction

Exercise-induced respiratory symptoms (EIS) are common in children and adolescents and most often present as dyspnea or shortness of breath, wheeze, and cough [1]. The most common causes of EIS are asthma leading to exercise-induced bronchoconstriction, insufficient fitness levels, and obesity [2,3]. In other cases, EIS may be a manifestation of an underlying pulmonary pathology such as dysfunctional breathing disorders or restrictive lung disease [3].

Childhood cancer survivors (CCS) represent a particularly vulnerable group for long-term pulmonary complications due to possible late effects of oncologic treatments [4]. Chest-directed radiotherapy, thoracic surgery, pulmotoxic chemotherapeutic agents (e.g., bleomycin, busulfan), and hematopoietic stem cell transplantation (HSCT) can lead to structural and functional lung damage, including interstitial lung disease and reduced pulmonary capacity [4,5]. While such impairments may remain asymptomatic at rest, they can become clinically evident during exercise, when respiratory demand increases [6].

Despite these known risks, EIS in CCS have been poorly characterized. Most studies reported on isolated symptoms, primarily wheeze [7] or dyspnea [8,9], rather than evaluating a broader spectrum of EIS. One study found a continued increase in the cumulative incidence of exercise-induced dyspnea over time since completion of treatment [9], which suggests a growing and persistent burden. However, the underlying causes of EIS in this population remain largely unexplored. Since EIS can contribute to avoiding physical activity [10,11], characterizing these symptoms in CCS is essential given the role physical activity can play in mitigating treatment-related late effects, supporting psychosocial reintegration, and improving cardiopulmonary health in survivors of childhood cancer [12,13]. We therefore investigated the prevalence of EIS, identified factors associated with EIS, and quantified the contributions of these factors to EIS using population attributable fractions.

## Methods

### Study design and population

We used cross-sectional data from the Swiss Childhood Cancer Survivor Study (SCCSS) FollowUp–Pulmo, a multicenter study of CCS attending pediatric oncology follow-up care in Switzerland. Detailed study procedures have been published previously [14]. Briefly, SCCSS FollowUp–Pulmo investigates lung health in CCS aged 6 to 20 years who had been diagnosed with cancer according to the International Classification of Childhood Cancer (ICCC-3) [15] at least one year before study entry. Eligible participants completed cancer treatment including systemic anticancer treatments (chemotherapy, immunotherapy, targeted agents), HSCT, thoracic surgery, or radiation to areas that may affect the lungs. We excluded CCS who were in a palliative or relapsed situation and those who were treated only with surgery or radiotherapy not affecting the chest area. The study collects clinical data from medical records and assesses symptoms and lifestyle via a questionnaire. For this analysis, we included participants enrolled between June 2022 and October 2024 from three pediatric oncology centers in Switzerland (Bern, Basel, and Geneva).

### Outcome: Exercise-induced symptoms

We obtained data on EIS from a questionnaire that included a set of questions on respiratory problems during exertion. For survivors younger than 14, parents completed the questionnaire, while those aged 14 years or older filled it in themselves. If respondents answered “yes” to the question “Does your child/Do you sometimes have breathing difficulties during exertion,” they were asked to specify the symptoms including dyspnea, cough, or wheeze. We coded EIS as a binary variable (yes/no) if participants reported at least one symptom. We also asked about activities during which EIS occurred (running short 50-100m or long >500m distances, cycling uphill, strenuous sports such as basketball, soccer, swimming, or other) to help differentiate potential underlying causes. These activities were selected based on their association with different mechanisms of EIS such as asthma, deconditioning, and obesity as previously described in the literature [16]. English translations of the original German/French questions are available in Supplementary Table S1.

### Explanatory variables

#### Sociodemographic and lifestyle characteristics

We retrieved information on participants’ age, height, and weight from medical records at the clinical visit corresponding to the date of questionnaire completion. Body mass index (BMI) was calculated and categorized according to World Health Organization (WHO) z-score standards: underweight (< –2 SD), normal weight (≥ –2 SD to ≤ +1 SD), overweight (> +1 SD), and obese (> +2 SD) [17]. Due to the small number of underweight participants, we combined them with the normal-weight group for analysis. To describe the population’s sociodemographic and behavioral context, we assessed parental education, migration background, and smoking behavior (active and passive) via the questionnaire. Migration background was considered positive if both parents were born outside of Switzerland, and negative if the participant and at least one parent were born in Switzerland. Parental education level was classified into three categories: primary (compulsory schooling only, ≤ 9 years), secondary (vocational training or upper secondary education), or tertiary (university or technical college education). Active smoking was assessed in CCS older than 14 years and passive smoking exposure in all participants. If participants engaged in recreational sports, they were asked to specify the type of sport, the number of hours per week spent on it, and if they had a medical condition limiting their physical activity. For further analysis, we classified participants as physically inactive if they did not report engaging in any recreational sports, presuming their reduced physical fitness level.

#### Cancer- and treatment-related characteristics

We extracted the following cancer- and treatment-related information from medical records: age at diagnosis, cancer diagnosis, time since diagnosis, chemotherapeutic agents, immune- and targeted therapies, HSCT (yes/no), thoracic surgery (yes/no), pulmotoxic radiotherapy (yes/no), and relapse status (yes/no). We defined pulmotoxic chemotherapy as having received busulfan, bleomycin, or nitrosoureas (carmustine, lomustine) [18]. Thoracic surgery included any procedure involving the lungs or chest wall, excluding central line placement [18]. Pulmotoxic radiotherapy was defined as radiation to the chest (mantle, mediastinal, or whole lung fields), abdomen (whole or any upper field), or total body irradiation [5,18]. We also extracted pulmonary diseases from medical records. Asthma was coded as “yes” if participants reported having asthma or using asthma medication in the questionnaire, or had a current diagnosis of asthma in their medical records.

### Statistical analysis

We used mean and standard deviation (SD) or median and interquartile range (IQR) for continuous variables and frequencies and percentages for categorical variables to describe sociodemographic and clinical characteristics. To identify factors associated with EIS, we performed a multivariable logistic regression. We selected a priori exposure variables based on clinical plausibility and existing literature [1,5,6], as conceptualized in Supplementary Figure S1. These included age, sex, BMI categories, time since diagnosis, treatment exposures (pulmotoxic chemotherapy, thoracic surgery, HSCT, and pulmotoxic radiotherapy), asthma, and physical inactivity. To estimate the potential impact of these factors, we calculated population attributable fractions (PAFs), using the *punaf* command in Stata [19]. PAFs were derived from the fully adjusted logistic regression model and represent the proportion of EIS cases that might be prevented if these exposures were eliminated or effectively managed at the population level, assuming a causal relationship [20,21]. We used Stata (Version 16.1, Stata Corporation, Austin, TX) for all analyses.

## Results

### Characteristics of the study population

Of 218 eligible survivors who attended pediatric oncology follow-up care in participating centers, 196 filled in the questionnaire (response rate 90%, Supplementary Figure S2). More than half (59%) were male (Table 1). Median age at the time of the study was 14 years (interquartile range [IQR] 10-17), median age at diagnosis was 5 years (IQR 3-9), and median time since diagnosis 7 years (IQR 4-10). The most common diagnoses were leukemia (52%), lymphoma (10%), and neuroblastoma (9%). Nineteen participants (10%) had received pulmotoxic chemotherapy, 25 (13%) pulmotoxic radiotherapy, 16 (8%) thoracic surgery, and 20 (10%) HSCT. Seventeen (8%) were obese. Twenty-two participants (11%) had asthma, and one was diagnosed with pulmonary graft-versus-host disease. CCS with EIS were older at study, older at diagnosis, engaged less in recreational sports, and more often had asthma compared to CCS without EIS.

**Table 1.** Characteristics of participating childhood cancer survivors with and without exercise-induced symptoms (N=196)

|  | All CCS<br>N=196 | CCS with EIS<br>N=46 | CCS without EIS<br>N=150 | P-value<sup>a</sup> |
| --- | --- | --- | --- | --- |
| **Sociodemographic and lifestyle characteristics** | **n (%)<sup>b</sup>** | **n (%)<sup>b</sup>** | **n (%)<sup>b</sup>** |  |
| Median age at study in years (IQR) | 14 (10–17) | 15 (12–18) | 13 (10–17) | 0.035 |
| Age groups |  |  |  | 0.496 |
| 6–10 | 46 (24) | 6 (13) | 40 (26) |  |
| 11–14 | 71 (36) | 16 (35) | 55 (37) |  |
| 15–20 | 79 (40) | 24 (52) | 55 (37) |  |
| Sex (female) | 81 (41) | 21 (46) | 60 (40) |  |
| BMI categories |  |  |  | 0.212 |
| Underweight | 9 (5) | 3 (7) | 6 (4) |  |
| Normal weight | 121 (62) | 23 (50) | 98 (65) |  |
| Overweight | 49 (25) | 14 (30) | 35 (23) |  |
| Obese | 17 (8) | 6 (13) | 11 (7) |  |
| Migration background <sup>c</sup> | 59 (30) | 10 (22) | 49 (33) | 0.158 |
| Parental education (highest level) <sup>c</sup> |  |  |  | 0.351 |
| Primary or secondary education | 82 (42) | 22 (48) | 60 (40) |  |
| Tertiary education | 102 (52) | 21 (46) | 81 (54) |  |
| Active smoking (n=92) <sup>e</sup> | 11 (12) | 3 (11) | 8 (12) | 0.590 |
| Passive smoking <sup>f</sup> | 59 (30) | 15 (33) | 44 (29) | 0.631 |
| Recreational sports | 145 (75) | 25 (56) | 120 (81) | 0.001 |
| **Cancer- and treatment-related characteristics** | **n (%)<sup>b</sup>** | **n (%)<sup>b</sup>** | **n (%)<sup>b</sup>** |  |
| Median age at diagnosis in years (IQR) | 5 (3–9) | 7 (3–13) | 4 (2–8) | 0.002 |
| Median time since diagnosis in years (IQR) | 7 (4–10) | 5 (4–9) | 7 (5–10) | 0.091 |
| Diagnosis (ICCC-3) |  |  |  | 0.384 |
| Leukemia | 102 (52) | 23 (50) | 79 (53) |  |
| Lymphoma | 20 (10) | 6 (13) | 14 (9) |  |
| CNS tumor | 11 (6) | 2 (4) | 9 (6) |  |
| Neuroblastoma | 18 (9) | 3 (7) | 15 (10) |  |
| Renal tumor | 15 (8) | 3 (7) | 12 (8) |  |
| Soft tissue sarcoma | 14 (7) | 2 (4) | 12 (8) |  |
| Other tumor <sup>g</sup> | 16 (8) | 7 (15) | 9 (6) |  |
| Treatment |  |  |  |  |
| Chemotherapy <sup>h</sup> | 194 (99) | 46 (100) | 148 (99) | 0.585 |
| Pulmotoxic <sup>i</sup> | 19 (10) | 4 (9) | 15 (10) | 0.526 |
| Pulmotoxic radiotherapy <sup>j</sup> | 25 (13) | 8 (18) | 17 (11) | 0.281 |
| Thoracic surgery <sup>k</sup> | 16 (8) | 5 (11) | 11 (7) | 0.443 |
| HSCT <sup>l</sup> | 20 (10) | 5 (11) | 15 (10) | 0.865 |
| Relapse | 15 (8) | 3 (7) | 12 (8) | 0.515 |
| **Pulmonary disease** | **n (%)<sup>b</sup>** | **n (%)<sup>b</sup>** | **n (%)<sup>b</sup>** |  |
| Asthma | 22 (11) | 14 (30) | 8 (5) | <0.001 |
| Pulmonary graft-versus-host disease | 1 (0.5) | 1 (2) | 0 (0) | 0.235 |

<sup>a</sup> P-value comparing childhood cancer survivors with and without exercise-induced symptoms derived from chi-squared or Fisher's exact test for categorical and t-test for continuous variables.
<sup>b</sup> Column percentages are given.
<sup>c</sup> Migration background was considered positive if both parents were born outside of Switzerland and negative if the participant and at least one parent were born in Switzerland.
<sup>d</sup> Data missing for 12 participants.
<sup>e</sup> Question on active smoking was only asked for participants 14 years of age or older, data of 5 participants are missing.
<sup>f</sup> Data missing for 11 participants.
<sup>g</sup> Including retinoblastoma, hepatic, bone, germ cell tumors, other malignant epithelial neoplasms, malignant melanomas, other or unspecified malignant neoplasms, and Langerhans cell histiocytosis.
<sup>h</sup> Any chemotherapy, targeted agents, or immunotherapy, alone or combined with other treatments.
<sup>i</sup> Including busulfan, bleomycin, and nitrosoureas.
<sup>j</sup> Radiotherapy involving chest, abdomen, spine, or total body irradiation alone or combined with other treatments.
<sup>k</sup> Surgery involving thorax or lungs alone or combined with other treatments.
<sup>l</sup> Any hematopoietic stem cell transplantation alone or combined with other treatments.
Abbreviations: CCS, childhood cancer survivors; EIS, exercise-induced symptoms; BMI, body mass index; IQR, interquartile range; CNS, central nervous system; ICC-3, International Classification of Childhood Cancer - Third Edition; HSCT, hematopoietic stem cell transplantation; NA, not applicable

While 51 CCS (26%, 95% confidence interval [CI] 20-33%) were physically inactive, most still engaged in recreational sports (74%, 95%CI: 68–81%) (Supplementary Table S2). On average, participants spent 4.2 hours per week on recreational sports. Types of recreational sports that CCS engaged in are presented in Supplementary Figure S3. Twenty-two participants (12%) reported having a medical condition that limited their physical activity, with musculoskeletal issues being the most common (Supplementary Table S2). Of those, 13 were physically inactive while 9 still engaged in recreational sports.

### Prevalence of exercise-induced symptoms

One out of every four of the 196 participants reported one or more EIS (24%, 95%CI: 18-30%). Dyspnea was reported most commonly, by 14% (95% CI 9-19%), followed by cough (12%, 95%CI: 8-18%), and wheezing (7%, 95%CI: 4-11%) (Table 2). Most participants reported symptoms when running long distances (>500m; 61%) or short distances (50-100m; 52%).

**Table 2.** Exercise-induced symptoms in childhood cancer survivors.

|  | N=196 |  |  |
| --- | --- | --- | --- |
|  | n | % | (95%CI) |
| **Exercise-induced symptoms<sup>a</sup>** | 46 | 24 | (18–30) |
| Dyspnea | 27 | 14 | (9–19) |
| Cough | 24 | 12 | (8–18) |
| Wheeze | 13 | 7 | (4–11) |
| **Activities during which symptoms occur<sup>b</sup>** | 46 |  |  |
| Running |  |  |  |
| Short distances (50-100m) | 24 | 52 | (37-67) |
| Long distances (>500m) | 28 | 61 | (45-75) |
| Cycling uphill | 18 | 39 | (25-55) |
| Strenuous exercise (e.g., soccer or basketball) | 21 | 46 | (31-61) |
| Swimming | 12 | 26 | (14-41) |
| Other<sup>c</sup> | 15 | 8 | (4-12) |

<sup>a</sup> Defined as having reported at least one of the respiratory symptoms (dyspnea, cough, or wheeze). Participants may have reported more than one symptom.
<sup>b</sup> Reported only by participants who had exercise-induced symptoms.
<sup>c</sup> Self-reported situations such as climbing upstairs or hiking.
Abbreviation: CI, confidence interval

### Factors associated with exercise-induced symptoms

Females (OR 2.5, 95%CI: 1.1-5.9), older survivors (odds ratio [OR] 1.2 per year of increase in age, 95%CI: 1.1-1.3), obese survivors (OR 4.7, 95%CI: 1.1-19.6, compared to normal weight), those with asthma (OR 10.1, 95%CI: 3.3-31.2), and physically inactive survivors (OR 2.3, 95%CI: 1.3-6.6) were more likely to experience EIS (Figure 1, Supplementary Table S3).

**Figure 1.**
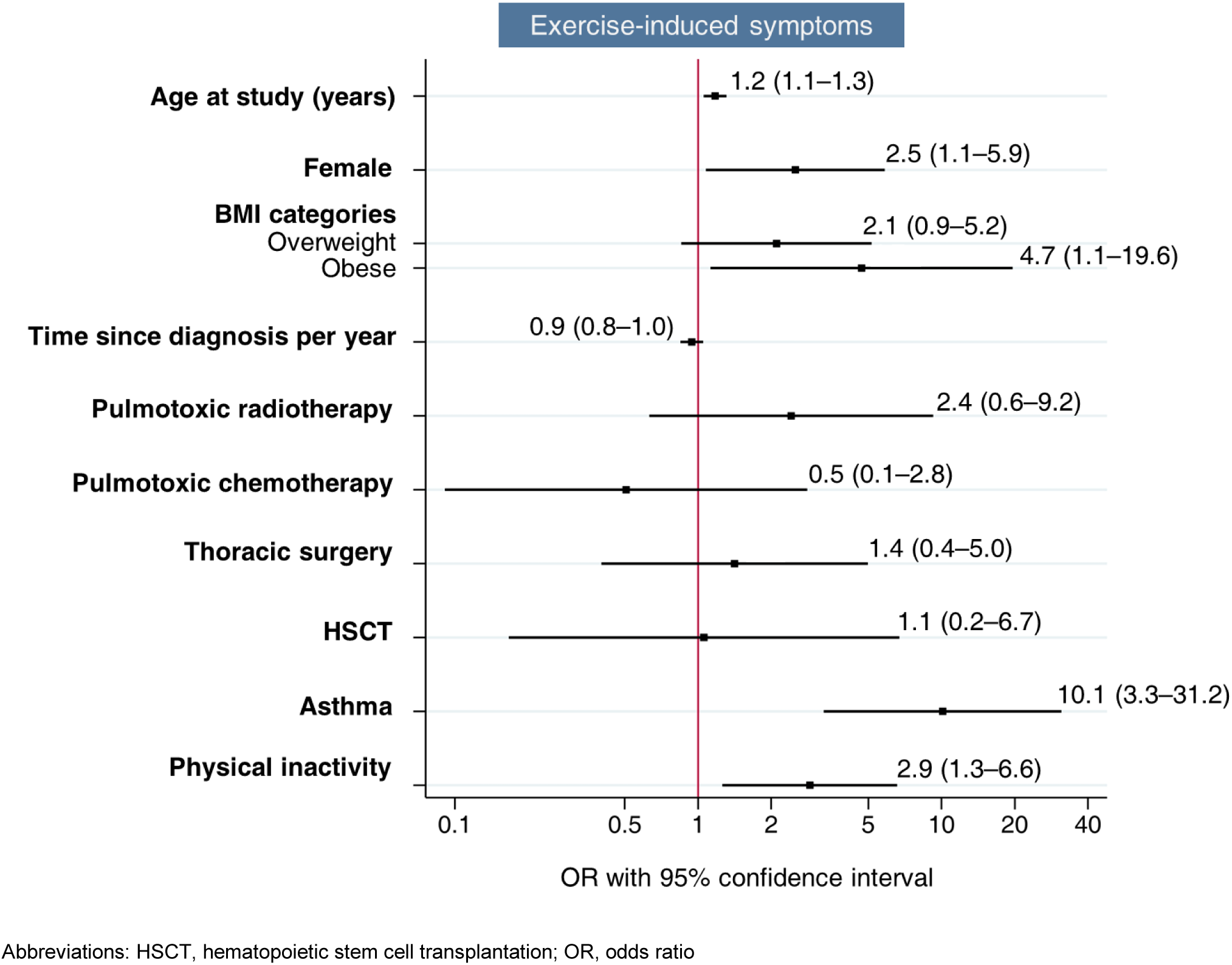
Factors associated with exercise-induced symptoms in childhood cancer survivors. Odds ratios from multivariable logistic regression adjusted for sociodemographic, lifestyle, and clinical characteristics.

Survivors who received pulmotoxic radiotherapy (OR 2.4, 95%CI: 0.6-9.2) showed a trend toward increased reporting of EIS. No associations were found with other treatment-related exposures. In adjusted analyses, an estimated 7% (95%CI: −0.5% to 15%) of EIS cases were attributable to obesity, 20% (95%CI: 9%–29%) to asthma, and 18% (95%CI: 2%–32%) to physical inactivity (Table 3). The joint PAF for obesity, asthma, and physical inactivity was 44% (95%CI: 24%–58%). PAFs for treatment-related exposures were small and had wide confidence intervals crossing zero.

**Table 3.** Estimated population attributable fractions and 95% confidence intervals for exposures associated with exercise-induced symptoms in childhood cancer survivors, based on adjusted logistic regression models.

| Exposure | PAF | 95%CI |
| --- | --- | --- |
| Overweight | 11% | -4 – 24% |
| Obesity | 7% | -0.5 – 15% |
| Pulmotoxic radiotherapy | 7% | -4 – 17% |
| Pulmotoxic chemotherapy | -4% | -16 – 6% |
| Thoracic surgery | 2% | -6 – 10% |
| HSCT | 0.3% | -12 – 11% |
| Asthma | 20% | 9 – 29% |
| Physical inactivity | 18% | 2 – 32% |

PAFs were calculated from the adjusted logistic regression model applying the approach by Greenland and Drescher [REF]. Abbreviations: PAF, population attributable fraction; CI, confidence interval

## Discussion

A substantial proportion of CCS—one in four—reported experiencing EIS. While survivors who received pulmotoxic radiotherapy showed a trend toward increased reporting EIS, exposure to cancer treatment was not conclusively associated with EIS. However, a higher likelihood of EIS was independently associated with female sex, older age, obesity, asthma, and physical inactivity. Notably among these, 44% of EIS could be attributed to obesity, asthma, and physical inactivity—risk factors that are modifiable or treatable with targeted interventions similar to those available to the general population.

Few previous studies have examined EIS in childhood cancer survivors and mainly focused on single symptoms. For example, a USA-based questionnaire study of predominantly adult CCS reported a prevalence of 11% for exercise-induced dyspnea [9], while a study in pediatric acute lymphoblastic leukemia survivors found exertional dyspnea in 15% [22]. In a Swiss nationwide cohort of CCS aged 5-16 years, 6% reported exercise-induced wheeze [7]. Our findings of 14% reporting dyspnea and 7% reporting wheeze are generally consistent with these previous studies. Comparative data from the general population are limited, as most studies report EIS in the context of asthma or other chronic respiratory conditions. Nonetheless, cross-sectional surveys have found similar prevalence rates: 8–12% for exertional wheeze in Swiss schoolchildren aged 6–17 years [23], 14% for exercise-induced dyspnea among Swedish adolescents aged 12–13 years [24], and 11% for exercise-induced cough in French primary school children [25]. While our sample size precludes detailed age-stratified comparisons, the frequency of individual symptoms in CCS appears largely comparable to that in peers without a cancer history. However, the overall burden of EIS may be underestimated when only individual symptoms are assessed. In our study, one in four CCS (24%) reported at least one EIS, substantially higher than the prevalence of any single symptom alone. This highlights the need to ask about different symptoms during follow-up care since focus on individual symptoms may underestimate the overall burden.

In our cohort, both asthma and obesity were significantly associated with EIS, which reflects patterns observed in the general pediatric population. Asthma is a well-established risk factor for EIS [6]. During physical exertion, increased ventilation cools and dries the airways, which can trigger bronchoconstriction and lead to symptoms such as wheezing, cough, and shortness of breath, particularly during prolonged aerobic activity [6,16]. This aligns with our observation that the most commonly reported triggers were long- and short-distance running, activities known to provoke symptoms in individuals with poorly controlled asthma or low cardiorespiratory fitness [16]. Notably, effective asthma management can enable children to remain physically active and symptom-free [26]. Obesity was also an important risk factor in our analysis and is known to impair respiratory mechanics through reductions in lung volumes, increased airway resistance, and greater work of breathing [27]. These physiological changes can contribute to the sensation of breathlessness during exercise, independent of underlying pulmonary disease [28]. In our population, an estimated 7% of EIS could be attributed to obesity and 20% to asthma. These findings emphasize the importance of recognizing and addressing these modifiable and treatable factors in both clinical assessments and survivorship care since targeted interventions could substantially reduce the symptom burden.

Physical inactivity was another major contributor to EIS in our cohort. While 74% of CCS reported participation in recreational sports, this was lower than the 82–88% participation rate observed among Swiss schoolchildren in a national survey [29], suggesting that survivors may be at increased risk of inactivity. Several factors may explain this: parents may adopt protective attitudes that discourage sports participation, and children may underestimate their own capabilities or feel discouraged by early fatigue or physical limitations [30,31]. Our findings nevertheless indicate that physical limitations are not always prohibitive; of the 12% of survivors who reported medical restrictions, 40% still engaged in sports. This suggests that with appropriate guidance and support, many CCS can remain active despite health challenges.

Importantly, we estimated that 18% of EIS in our study could be attributed to physical inactivity, and the joint PAF for asthma, obesity, and inactivity was 44%. This means that nearly half of EIS cases could potentially be prevented or mitigated by addressing non-cancer-related factors. The relationship between physical activity and EIS is likely bidirectional: symptoms may discourage exercise, while deconditioning from inactivity may worsen symptoms. However, evidence from the general population suggests that regular physical activity can reduce EIS, possibly by improving airway function and reducing inflammation [32]. Taken together, these results emphasize the potential for low-risk, modifiable interventions—including asthma management, weight control, and tailored exercise programs—to reduce EIS and improve overall health outcomes in CCS.

Sex and age, while not modifiable, may help identify CCS at higher risk for EIS. In our cohort, females were more likely than males to report EIS, which is consistent with findings from studies in healthy populations [33,34]. This may reflect anatomical differences since females generally have smaller airways and lower lung volumes relative to body size, which increases the work of breathing and the likelihood of expiratory flow limitation during exertion [35]. We also found that older age was associated with higher EIS prevalence, which is in line with previous research suggesting that adolescents may engage in more strenuous physical activity and be more aware of their symptoms [36,37]. These findings suggest that physiological and perceptual factors may contribute to symptom burden and should be considered in risk stratification and individualized follow-up care.

Regarding treatment-related factors, our study did not find clear associations between pulmotoxic therapies and EIS. However, there was a tendency toward increased risk among CCS who received pulmotoxic radiotherapy. This aligns with findings from a study in adult CCS in which chest-directed radiotherapy was associated with higher rates of exercise-induced dyspnea, and the cumulative incidence continued to rise even decades after treatment [9].

Radiation-induced lung injury can lead to progressive parenchymal and interstitial changes, with studies suggesting these effects may worsen with age even if they remain clinically silent during childhood or adolescence [38,39]. In our cohort, the median time since diagnosis was 7 years, which may be insufficient for these late effects to become fully apparent. Additionally, small numbers of survivors exposed to specific treatments may have limited our ability to detect statistically significant associations. Future research should include larger, longitudinal studies with long follow-up to better characterize the long-term effect of cancer therapies on EIS.

A key strength of our study is its high participation rate (90%) supporting the generalizability of our findings to CCS with similar cancer and treatment profiles. By assessing multiple EIS, we captured a more comprehensive picture of symptom prevalence than studies focusing on single symptoms. However, the cross-sectional design limits causal inference, and reliance on self-reported data may introduce recall or reporting bias. Still, self-reporting remains valuable since it captures participants’ subjective experiences that are clinically relevant. The relatively small number of EIS cases limited statistical power for subgroup analyses by treatment or diagnosis. Although we identified several modifiable contributors to EIS, nearly half of the symptom burden remained unexplained. Future research should explore additional factors such as undiagnosed cardiopulmonary disease, neuromuscular weakness, and psychological contributors like fatigue and anxiety, which may influence symptom perception. Objective assessments, such as cardiopulmonary exercise testing, could help clarify underlying mechanisms and guide targeted interventions.

In summary, we found that a median 7 years after cancer diagnosis EIS affect a substantial proportion of CCS, and in our cohort nearly half of these symptoms could be attributed to modifiable or treatable risk factors including asthma, obesity, and physical inactivity. Clinicians should be aware of these symptoms and first assess common, treatable causes before attributing them to late effects of cancer treatment. Early identification and targeted intervention for modifiable contributors could help reduce symptom burden, support physical activity, and improve long-term health outcomes in CCS.

## Statements & Declarations

## Acknowledgements

We thank all survivors for participating in our study, Pediatric Hematology/Oncology and Pediatric Pulmonology teams at the University Hospitals in Bern, Basel, and Geneva, and the study team of the Childhood Cancer Research Group. We also thank Christopher Ritter for editorial assistance.

## Author contribution

Maša Žarković: formal analysis, writing—original draft preparation, visualization; Carina Nigg: formal analysis, writing—review and editing; Christina Schindera: writing—review and editing; Myrofora Goutaki: writing—review and editing; Sonja Lüer: writing—review and editing; Marc Ansari: writing—review and editing; Claudia E Kuehni: conceptualization, methodology, writing— review and editing, funding acquisition, supervision.

## Data availability

Researchers interested in collaborative work can contact the corresponding author (Claudia Kuehni) to discuss planned projects or analyses of existing data.

## Ethics approval

The Ethics Committee of the Canton of Bern (2019-00739) granted ethical approval.

## Consent to participate

All participants or their guardians provided informed consent.

---

**Supplementary Figure S1.**
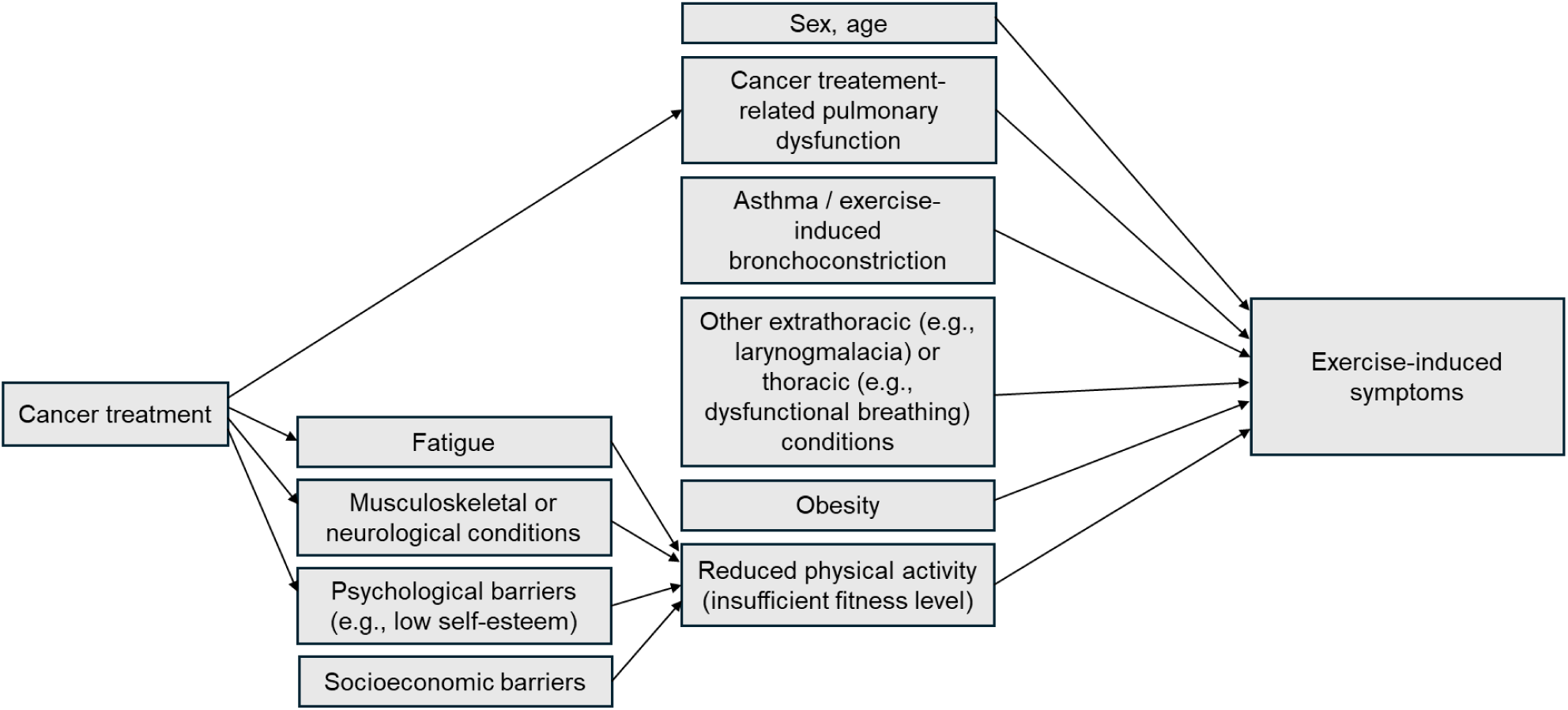
Conceptual framework showing possible predictors of exercise-induced symptoms and reduced physical activity as related to cancer treatments.

**Supplementary Figure S2.**
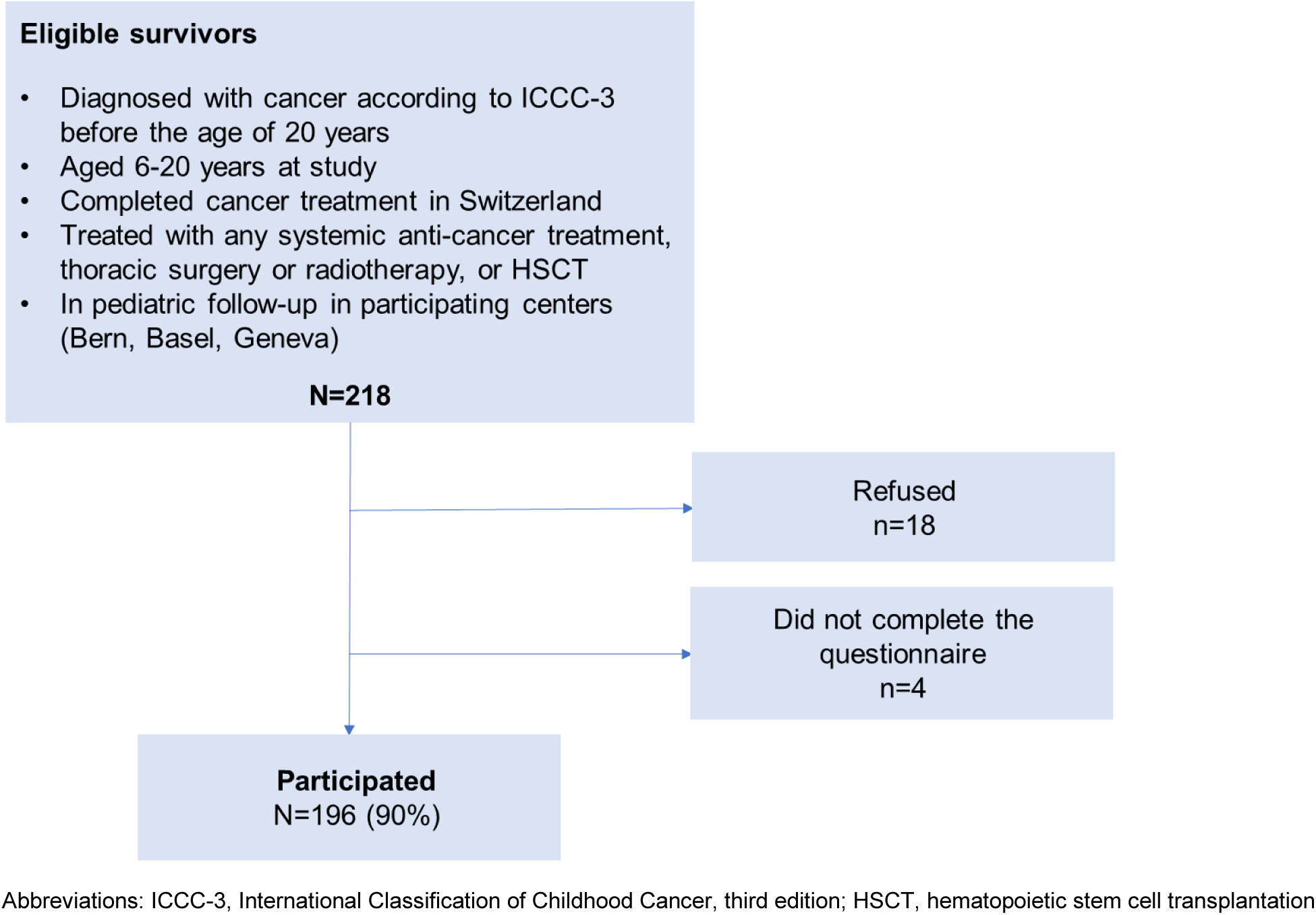
Flow diagram of the study population.

**Supplementary Figure S3.**
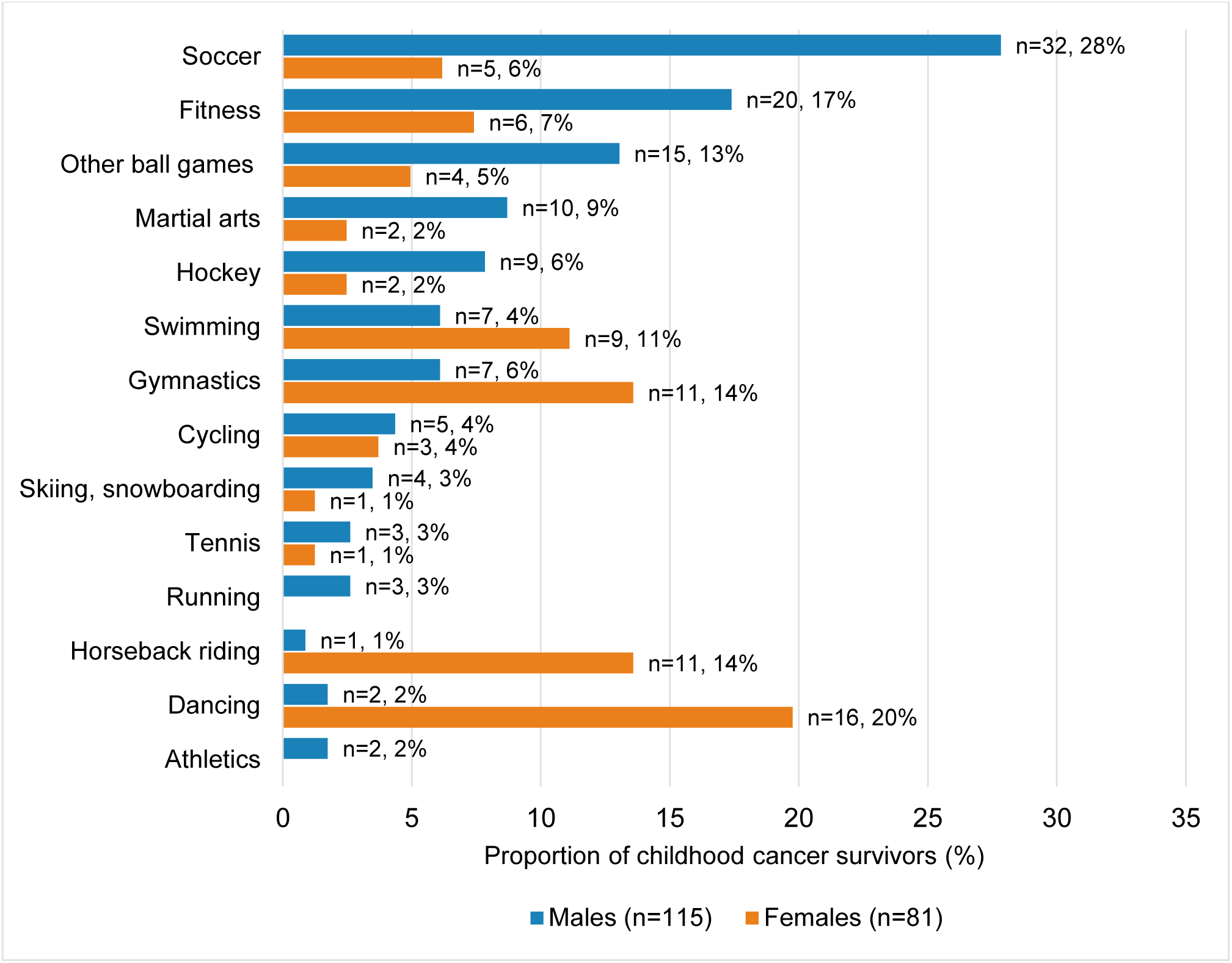
Recreational sport types and frequencies among childhood cancer survivors stratified by sex. There can be multiple (1-3) recreational sport types per child.

**Supplementary Table S1.**
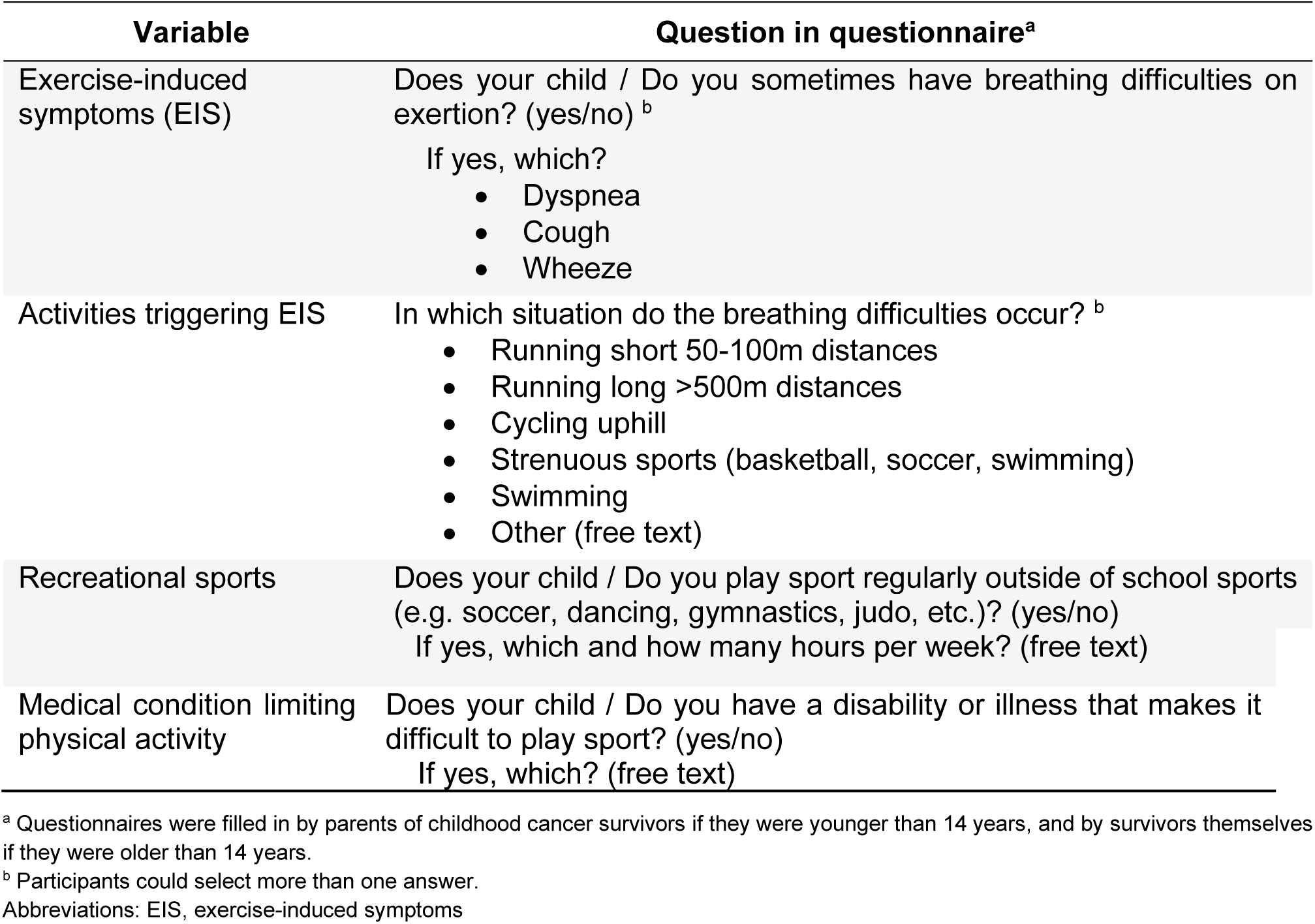
English translations of original German/French questionnaire items on exercise-induced symptoms and recreational sports.

**Supplementary Table S2.** Prevalence of recreational sport engagement and physical inactivity across age groups, and self-reported physical activity limitations.

|  | N=196 |  |  |
| --- | --- | --- | --- |
|  | n | % | (95%CI) |
| **Recreational sports** | 145 | 74<sup>a</sup> | (68–81) |
| *Age categories, years* |  |  |  |
| 6-10 | 35 | 76<sup>b</sup> | (61–87) |
| 11-14 | 58 | 82<sup>b</sup> | (71–90) |
| 15-20 | 52 | 66<sup>b</sup> | (55–77) |
| *Average hours/week of recreational sports, mean, SD* |  | 4.2 (2.9) |  |
| **Physical inactivity** | 51 | 26<sup>b</sup> | (20–33) |
| *Age categories, years* |  |  |  |
| 6-10 | 11 | 24<sup>b</sup> | (13–39) |
| 11-14 | 13 | 18<sup>b</sup> | (10–29) |
| 15-20 | 27 | 34<sup>b</sup> | (24–46) |
| **Physical activity limitations <sup>c,d</sup>** | 22 | 11<sup>a</sup> | (7–17) |
| Musculoskeletal conditions <sup>e</sup> | 12 | 6<sup>a</sup> | (3–10) |
| Cardiorespiratory conditions <sup>f</sup> | 3 | 2<sup>a</sup> | (0–4) |
| Neurological conditions <sup>g</sup> | 3 | 2<sup>a</sup> | (0–4) |
| Other <sup>h</sup> | 4 | 2<sup>a</sup> | (0–5) |

<sup>a</sup> Column percentages are presented.
<sup>b</sup> Row percentages are presented.
<sup>c</sup> Of 22 participants who reported physical activity limitations, 9 (41%) engaged in recreational sports.
<sup>d</sup> Reporting a medical condition perceived as limiting physical activity.
<sup>e</sup> Self-reported musculoskeletal conditions such as joint necrosis, shorter or missing leg, or using a wheelchair.
<sup>f</sup> Self-reported cardiorespiratory conditions such as asthma or heart rhythm disorder.
<sup>g</sup> Self-reported neurological conditions such as epilepsy.
<sup>h</sup> Self-reported conditions such as unspecified problems after cancer disease.

**Supplementary Table S3.**
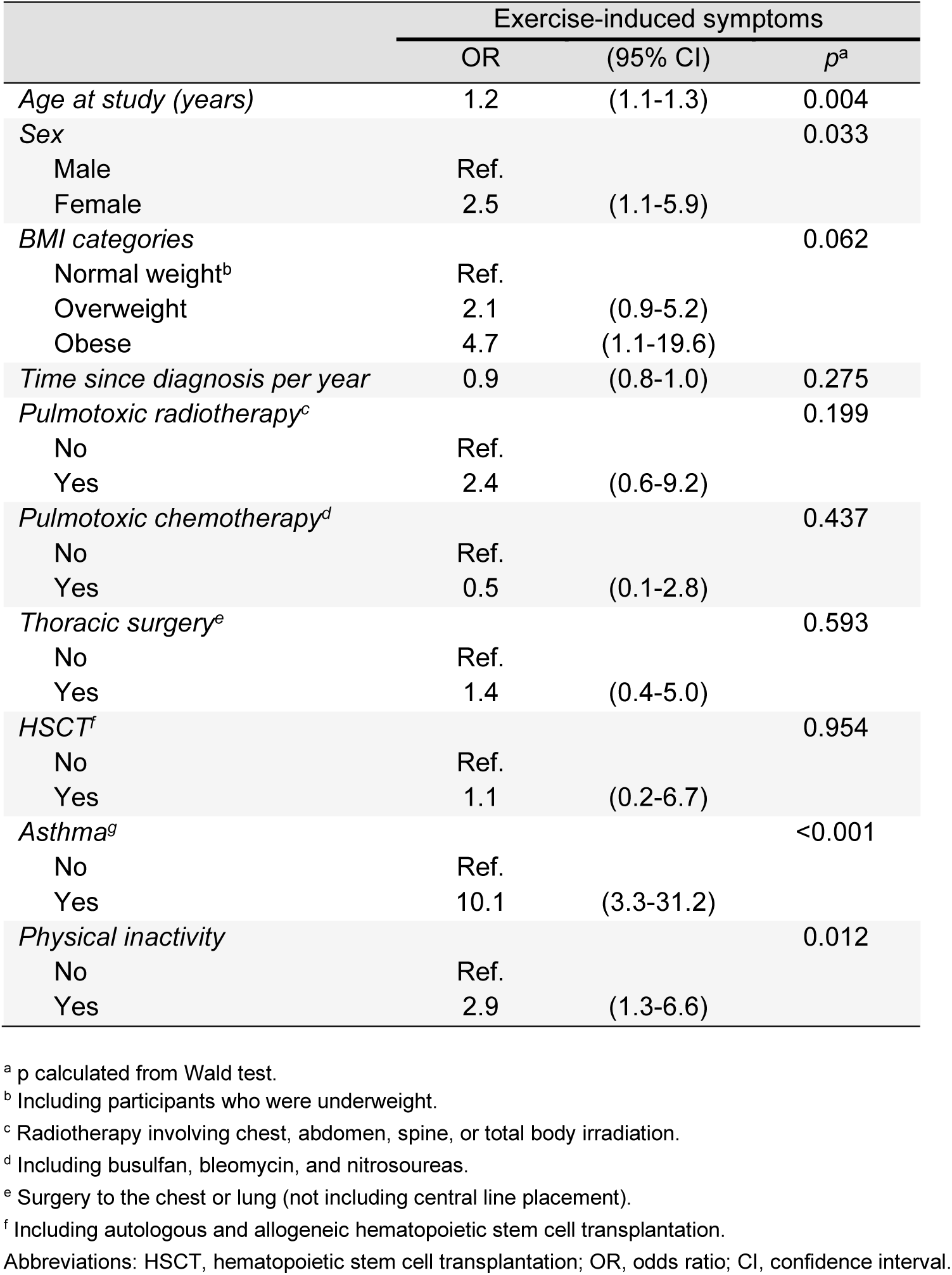
Association between exercise-induced symptoms and sociodemographic, lifestyle, and clinical characteristics in childhood cancer survivors using multivariable logistic regression.

## References

1 Pedersen ESL, Ardura-Garcia C, de Jong CCM, et al. Diagnosis in children with exercise-induced respiratory symptoms: A multi-center study. Pediatr Pulmonol. 2021;56:217–25. doi: 10.1002/PPUL.25126

2 Malmberg M, Malmberg LP, Pelkonen AS, et al. Overweight and exercise-induced bronchoconstriction – Is there a link? Pediatric Allergy and Immunology. 2021;32:992–8. doi: 10.1111/pai.13492

3 Grandinetti R, Mussi N, Rossi A, et al. Exercise-Induced Bronchoconstriction in Children: State of the Art from Diagnosis to Treatment. J Clin Med. 2024;13.

4 Huang T-T, Hudson MM, Stokes DC, et al. Pulmonary Outcomes in Survivors of Childhood Cancer. Chest. 2011;140:881–901. doi: 10.1378/chest.10-2133

5 Visscher H, Otth M, Feijen EAM (Lieke), et al. Cardiovascular and Pulmonary Challenges After Treatment of Childhood Cancer. Pediatr Clin North Am. 2020;67:1155–70.

6 Del Giacco SR, Firinu D, Bjermer L, et al. Exercise and asthma: an overview. Eur Clin Respir J. 2015;2:27984. doi: 10.3402/ecrj.v2.27984

7 Žarković M, Sommer G, Nigg C, et al. Parental smoking and respiratory outcomes in young childhood cancer survivors. Pediatr Blood Cancer. 2024;71. doi: 10.1002/pbc.31310

8 Record E, Williamson R, Wasilewski-Masker K, et al. Analysis of Risk Factors for Abnormal Pulmonary Function in Pediatric Cancer Survivors. Pediatr Blood Cancer. 2016;63:1264–71. doi: 10.1002/pbc.25969

9 Mertens AC, Yasui Y, Liu Y, et al. Pulmonary complications in survivors of childhood and adolescent cancer: A report from the Childhood Cancer Survivor Study. Cancer. 2002;95:2431–41. doi: 10.1002/cncr.10978

10 Brasholt M, Baty F, Bisgaard H. Physical activity in young children is reduced with increasing bronchial responsiveness. Journal of Allergy and Clinical Immunology. 2010;125:1007–12. doi: 10.1016/j.jaci.2010.02.002

11 Glazebrook C, McPherson AC, Macdonald IA, et al. Asthma as a Barrier to Children’s Physical Activity: Implications for Body Mass Index and Mental Health. Pediatrics. 2006;118:2443–9. doi: 10.1542/peds.2006-1846

12 Antwi GO, Jayawardene W, Lohrmann DK, et al. Physical activity and fitness among pediatric cancer survivors: a meta-analysis of observational studies. Supportive Care in Cancer. 2019;27:3183–94. doi: 10.1007/s00520-019-04788-z

13 Wogksch MD, Goodenough CG, Finch ER, et al. Physical activity and fitness in childhood cancer survivors: A scoping review. Aging Cancer. 2021;2:112–28. doi: 10.1002/aac2.12042

14 Žarković M, Schindera C, Sommer G, et al. Assessing Pulmonary Function in Children and Adolescents After Cancer Treatment: Protocol for a Multicenter Cohort Study (Swiss Childhood Cancer Survivor Study FollowUp–Pulmo). JMIR Res Protoc. 2025;14:e69743. doi: 10.2196/69743

15 Steliarova-Foucher E, Stiller C, Lacour B, et al. International classification of childhood cancer, third edition. Cancer. 2005;103:1457–67. doi: 10.1002/cncr.20910

16 Pedersen ESL, de Jong CCM, Ardura-Garcia C, et al. Reported Symptoms Differentiate Diagnoses in Children with Exercise-Induced Respiratory Problems: Findings from the Swiss Paediatric Airway Cohort (SPAC). J Allergy Clin Immunol Pract. 2021;9:881–889.e3. doi: 10.1016/j.jaip.2020.09.012

17 de Onis M. Development of a WHO growth reference for school-aged children and adolescents. Bull World Health Organ. 2007;85:660–7. doi: 10.2471/BLT.07.043497

18. Children’s Oncology Group. Long-Term Follow-Up Guidelines for Survivors of Childhood, Adolescent, and Young Adult Cancers.

19 Newson RB. Attributable and Unattributable Risks and Fractions and other Scenario Comparisons. The Stata Journal: Promoting communications on statistics and Stata. 2013;13:672–98. doi: 10.1177/1536867X1301300402

20 Greenland S, Drescher K. Maximum likelihood estimation of the attributable fraction from logistic models. Biometrics. 1993;49:865–72.

21 Rückinger S, von Kries R, Toschke AM. An illustration of and programs estimating attributable fractions in large scale surveys considering multiple risk factors. BMC Med Res Methodol. 2009;9:7. doi: 10.1186/1471-2288-9-7

22 Arpaci T, Kilicarslan Toruner E. Assessment of problems and symptoms in survivors of childhood acute lymphoblastic leukaemia. Eur J Cancer Care (Engl). 2016;25:1034–43. doi: 10.1111/ecc.12561

23 Mozun R, Ardura-Garcia C, Pedersen ESL, et al. Agreement of parent- and child-reported wheeze and its association with measurable asthma traits. Pediatr Pulmonol. 2021;56:3813–21. doi: 10.1002/ppul.25690

24 Johansson H, Norlander K, Malinovschi A. Increased prevalence of exercise-induced airway symptoms – A five-year follow-up from adolescence to young adulthood. Respir Med. 2019;154:76–81. doi: 10.1016/j.rmed.2019.06.011

25 Momas I, Dartiguenave C, Fauroux B, et al. Prevalence of asthma or respiratory symptoms among children attending primary schools in Paris. Pediatr Pulmonol. 1998;26:106–12. doi: 10.1002/(SICI)1099-0496(199808)26:2<106::AID-PPUL6>3.0.CO;2-K

26 Vahlkvist S, Inman MD, Pedersen S. Effect of asthma treatment on fitness, daily activity and body composition in children with asthma. Allergy. 2010;65:1464–71. doi: 10.1111/j.1398-9995.2010.02406.x

27 Winck AD, Heinzmann-Filho JP, Soares RB, et al. Effects of obesity on lung volume and capacity in children and adolescents: a systematic review. Revista Paulista de Pediatria (English Edition). 2016;34:510–7. doi: 10.1016/j.rppede.2016.03.013

28 Shim YM, Burnette A, Lucas S, et al. Physical Deconditioning as a Cause of Breathlessness among Obese Adolescents with a Diagnosis of Asthma. PLoS One. 2013;8:e61022. doi: 10.1371/journal.pone.0061022

29. Lamprecht M, Bürgi R, Gebert A, et al. Sport Schweiz 2020 – Kinder- und Jugendbericht. 2021.

30 Bertorello N, Manicone R, Galletto C, et al. Physical Activity and Late Effects in Childhood Acute Lymphoblastic Leukemia Long-Term Survivors. Pediatr Hematol Oncol. 2011;28:354–63. doi: 10.3109/08880018.2010.550987

31 Larsen EH, Mellblom AV, Larsen MH, et al. Perceived barriers and facilitators to physical activity in childhood cancer survivors and their parents: A large-scale interview study from the International PACCS Study. Pediatr Blood Cancer. 2023;70. doi: 10.1002/pbc.30056

32 Mahler DA. Is physical activity anti-inflammatory on the airways? Thorax. 2007;62:376–376. doi: 10.1136/thx.2006.074021

33 Movahed MR, Martinez A, Morrell H, et al. Differences according to gender in reporting physical symptoms during echocardiographic screening in healthy teenage athletes. Cardiol Young. 2008;18:303–6. doi: 10.1017/S104795110800214X

34 Johansson H, Norlander K, Hedenström H, et al. Exercise-induced dyspnea is a problem among the general adolescent population. Respir Med. 2014;108:852–8. doi: 10.1016/j.rmed.2014.03.010

35 Dominelli PB, Molgat-Seon Y. Sex, gender and the pulmonary physiology of exercise. Eur Respir Rev. 2022;31. doi: 10.1183/16000617.0074-2021

36 Jurca M, Ramette A, Dogaru CM, et al. Prevalence of cough throughout childhood: A cohort study. PLoS One. 2017;12:e0177485. doi: 10.1371/journal.pone.0177485

37 Johansson H, Emtner M, Janson C, et al. The course of specific self-reported exercise-induced airway symptoms in adolescents with and without asthma. ERJ Open Res. 2020;6:00349–2020. doi: 10.1183/23120541.00349-2020

38 De A, Mascarenhas L, Kamath S, et al. Pilot feasibility study of comprehensive pulmonary evaluation following lung radiation therapy. J Pediatr Hematol Oncol. 2015;37:e412–8. doi: 10.1097/MPH.0000000000000407

39 Armstrong GT, Kawashima T, Leisenring W, et al. Aging and Risk of Severe, Disabling, Life-Threatening, and Fatal Events in the Childhood Cancer Survivor Study. Journal of Clinical Oncology. 2014;32:1218–27. doi: 10.1200/JCO.2013.51.1055

